# Identification Cure Hub Genes of Chromophobe Cell Renal Carcinoma : A study based on Weighted Gene Co-expression Network Analysis (WGCNA) and the Cure Defective Models

**DOI:** 10.1101/2023.07.01.23292107

**Authors:** Maryam Ahmadian, Zahra Molavi, Ahmad Reza Baghestani, Ali Akbar Maboudi

## Abstract

Renal cell carcinoma (RCC) is a prevalent and aggressive tumor of the urinary system with limited treatment success and poor patient outcomes. However, some patients exhibit long-term symptom relief and are considered ’cured’ after successful treatment. This study explores the genetic and pathway mechanisms underlying RCC cure for the first time, utilizing a survival model called the 3-parameter defective Gompertz cure model.

The study methodology involved two main steps: Firstly, employing Weighted Gene Co-expression Network Analysis (WGCNA) for gene network analysis, which identified six key modules associated with different aspects of cancer progression and survival. Hub genes, pivotal in cellular interactions, were pinpointed through network analysis. Secondly, the 3-parameter defective Gompertz model was utilized to identify therapeutic genes linked to successful treatment outcomes (CSRGs) in RCC. These genes were then compared with genes associated with patient survival (SRGs) using a cox model.

The study found ten hub genes commonly identified by both the defective 3-parameter Gompertz and Cox models, with six genes (NCAPG, TTK, DLGAP5, TOP2A, BUB1B, and BUB1) showing strong predictive values. Moreover, six hub genes (TTK, KIF20A, DLGAP5, BUB1, AURKB, and CDC45) were highlighted by the defective Gompertz model as significantly impacting cure when expressed at high levels. Targeting these hub genes may hold promise for improving RCC treatment outcomes and prognosis prediction.

Overall, this study provides valuable insights into the molecular mechanisms of RCC and underscores the potential of the defective 3-parameter Gompertz model in guiding targeted therapeutic approaches.

## 1. Introduction

Renal cell carcinoma (RCC) is a type of cancer that accounts for 5% and 3% of all oncological diagnoses worldwide in men and women, respectively (Capitanio et al., 2019). Men are almost twice as likely to develop kidney cancer as women, and the incidence rates of RCC increase consistently with advancing years, reaching a peak at about age 75(Scelo and Larose, 2018). About half of all RCC cases are identified before the age of 65(Huang et al., 2021a, Capitanio et al., 2019). RCC is classified into four pathological phases based on tumor size, invasion extent, and metastasis. Most RCCs arise in the kidney’s cortex, including the tubular apparatus, glomerulus, and collecting duct(Padala et al., 2020).

There are numerous genetic, clinical, and environmental risk factors associated with RCC, including smoking, drinking, high blood pressure, obesity, and occupational and environmental exposures to toxins such as cadmium, asbestos, and ionizing radiation. Inherited disorders, including Von Hippel-Lindau disease, hereditary leiomyomata’s RCC, and hereditary papillary RCC, have also been linked to renal carcinoma (Chow et al., 2010, Sims et al., 2018, Pastore et al., 2015). The cure rate for kidney cancer depends on various factors such as the stage of cancer, age, and treatments used. Early detection and prompt treatment lead to a high cure rate, with a five-year survival rate of 93% for stage I kidney cancer. However, if the cancer has spread, the cure rate drops significantly, with a five-year survival rate of around 12% for stage IV kidney cancer(Pandey and Syed, 2020, Sung et al., 2018).

The identification of biomarkers and potential therapeutic targets for RCC progression is a crucial area of research in cancer biology. In this study, the main focus was on identifying cured survival-related genes (CSRGs) in RCC using defective Gompertz 3-parameter model and compare result achieved by survival-related genes (SRGs) with common method of Cox regression has been used to found SRGs. In the standard survival models assume that all subjects are susceptible to the event of interest (such as recurrence or death from the disease). Cox regression is a standard survival model and a statistical technique used to examine how multiple variables influence the time until a specific event occurs. This method is considered semi-parametric since it does not make assumptions about the exact distribution of the data(Ihwah, 2015, Wulandari et al., 2021). The unsusceptible individuals that respond favorably to treatment appear to be free of symptoms in long-term follow up and may be regarded as ‘cured’ or immune and these individuals will never experience the event of interest; these risk-free subjects. Cox method cannot survey this characteristic of subjects. So, we need to kind of models that able to monitoring the immune individuals.

Models based on defective distributions use to estimate cure model. These distributions permit us to fit survival data consist of both cured and susceptible individuals. The proportion of the cured population is determined by computing the limit of the survival function defective distributions (cure rate that shown as parameter P) which range from zero to one (Balka et al., 2009, Masud et al., 2018).

In Kaplan-Meier curve, existence of the cured fraction is indicated by a long flat tail which is not close to zero (Rondeau, 2010); as it is shown in Figure 1.

**Fig.1.**
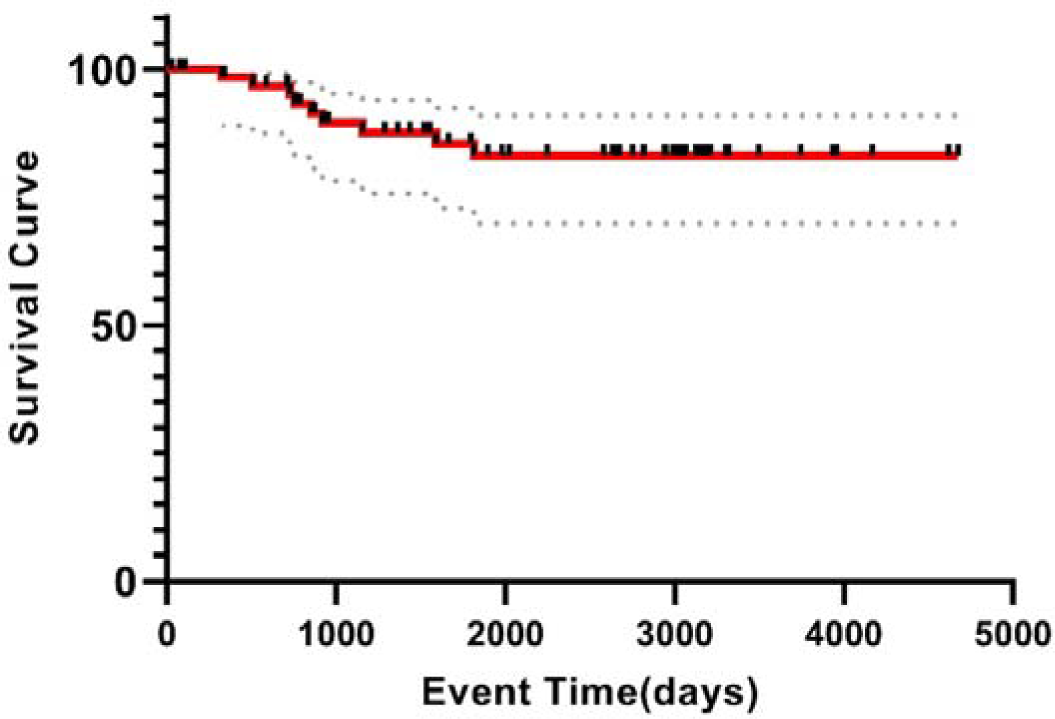
Kaplan-Meier curve of the kidney chromophobe cancer in TCGA data.

In this research we used defective 3-parameter Gompertz model for estimating cure rate. It’s proposed by Haji Zadeh and Baghestani (2022). It examines the impact of variables that resulted in risk-free subjects and cure rate (Hajizadeh et al., 2023). This model is important because of the majority of studies have focused on identifying SRGs and hazard disease-related gene combinations, while CSRGs have been largely ignored. this issue is a reason for difference between the common method of cox and the defective 3-parameter Gompertz model. It could be applied to kidney cancer data to gain a better understanding of its effectiveness.

By incorporating this model into the analysis, researchers can potentially identify new biomarkers and therapeutic targets for kidney cancer treatment. In this study, for the first time, examines the genes that effected on cure of patients and this issue has not been investigated in previous studies.

In summary, we applied WGCNA to the TCGA-KICH dataset to identify gene modules associated with survival outcomes in kidney cancer patients. Furthermore, both Cox proportional hazards regression and the defective Gompertz 3-parameters model were utilized to identify potential cures based on gene expression levels within these modules (figure 2).

**Fig.2.**
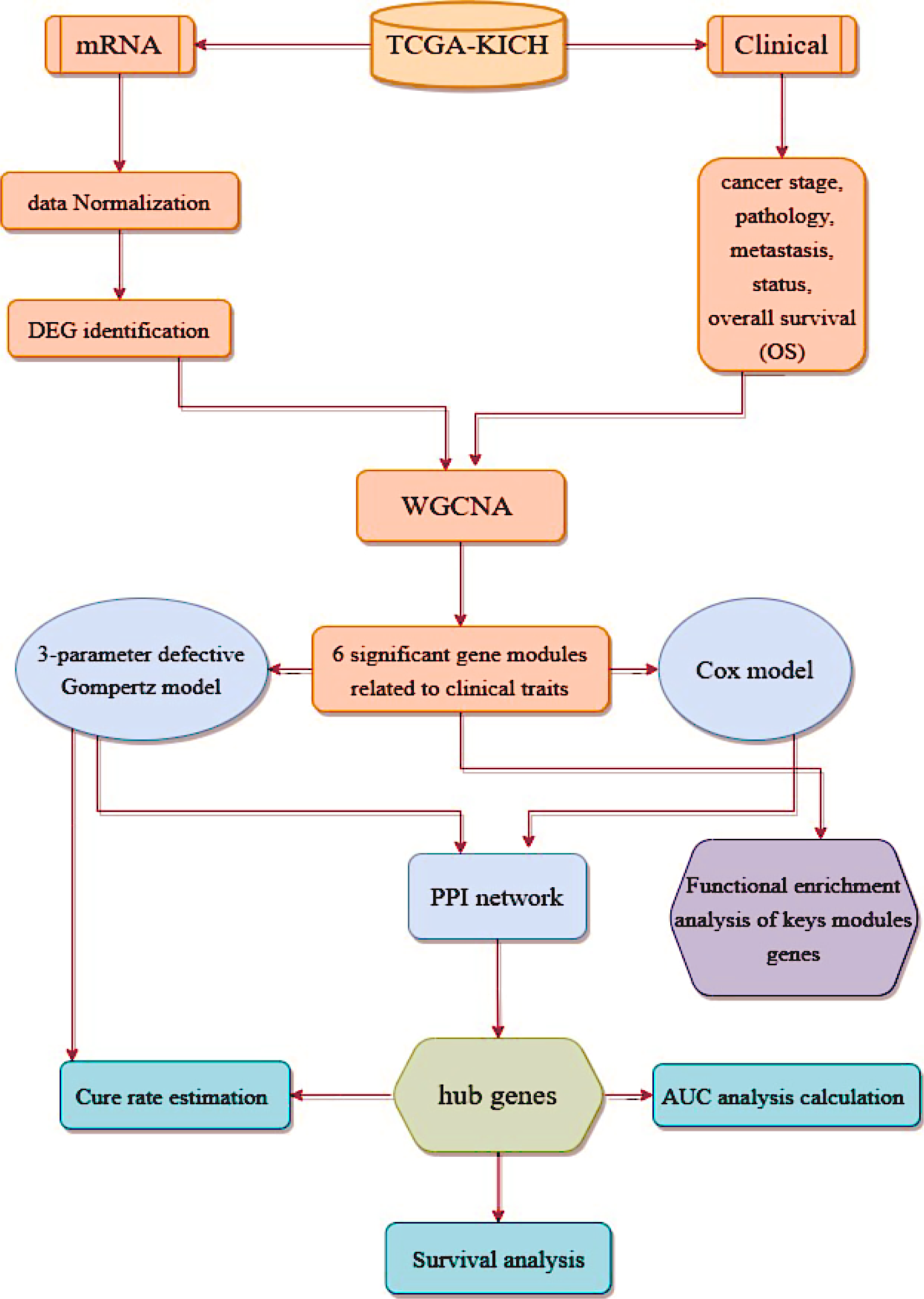
Flow chart of data preparation, processing, analysis, and validation.

## 2. Methods

### 2.1. Data selection

RNA-seq data and clinical information from 90 patients with kidney cancer were collected from Cancer Genome Atlas (TCGA) database (https://portal.gdc.cancer.gov/). The RNA-seq data consisted of 25 normal cases and 65 cancer cases. The clinical information included data on the overall survival (OS) of patients with kidney cancer.

### 2.2. KICH differentially expressed genes (DEGs) identification

To reduce the impact of outliers on the overall data, 7940 genes were selected based on an average expression greater than 1 and the greatest MAD. The Limma R software package was utilized for identifying DEGs between different groups. First, the ’lmFit’ function was used for multiple linear regression analysis on the expression spectrum dataset. The significance of DEGs was determined using the ’eBays’ function. A sample cluster map was drawn using the R language to identify and remove any outliers. Finally, the gene expression profiles were normalized using log(x+1) transformation to ensure that upregulated or downregulated genes were evenly distributed around 0. Volcano plot of the DEGs were created using the ’ggplot2’ function in R.

### 2.3. Calculation of gene Co-expressing using WGCNA

The process of calculating the Co-expression between different genes involved in kidney cancer were performed using the Weighted Gene Co-expression Network Analysis (WGCNA) package. This approach involves constructing a network of genes based on their expression patterns and then measuring the similarity or “correlation” between the genes in the network. It is a powerful tool for uncovering the complex interactions between genes and for identifying key players in the disease process(Langfelder and Horvath, 2007). The DEG data screened out in the TCGA KICH data were selected to establish a weighted gene co-expression network with WGCNA package(Zhao et al., 2010) in R Studio 3.6.0 software (http://www.r-project.org).

Then, a weighted adjacency matrix was constructed using a power function *A_mn_*[=|*C_mn_*|^β^ (*C_mn_*[=[Pearson’s correlation between genes *m* and *n*; *A_mn_*[=[adjacency between genes *m* and *n*), and a soft-thresholding parameter was applied to emphasize strong correlations (β) while penalizing weak ones. After selecting the power parameter β, the adjacency matrix was converted into a topological overlap matrix (Rocha et al.), which measures the network connectivity of a gene as the sum of its adjacency with all other genes in the network. The corresponding dissimilarity (1-TOM) was then calculated. To cluster genes with similar expression profiles into dissimilarity (1-TOM) was then calculated. To cluster genes with similar expression profiles into gene modules, average linkage hierarchical clustering was performed using the TOM-based dissimilarity measure. Gene dendrograms were constructed with a minimum size of 30 for gene groups. The module eigengenes dissimilarity measure was used to further analyze the gene modules. The cut line of the module dendrogram was selected and then some modules were merged to refine the analysis (Langfelder and Horvath, 2008, Ravasz et al., 2002, Yip and Horvath, 2007).

### 2.4. Identifying significant gene modules related to clinical traits

The WGCNA algorithm employs the module eigengene (ME) concept to assess the relationship between gene modules and clinical traits. ME is determined as the principal component obtained through a principal component analysis that captures the expression pattern of genes within a specific module. To identify the module highly correlated with RCC, the Pearson correlation coefficient between ME and clinical traits was computed. For intramodular analysis, gene significance (Masud et al.) and module membership (MM) were calculated. MM represents the correlation between ME and the gene expression profile and GS are the logarithmic transformation (log10) of the P-value (GS = lgP) indicating the correlation between gene expression and the clinical trait.

### 2.5. Functional enrichment analysis of keys module genes

To further understand the function of the selected genes, a functional enrichment analysis was performed using the online database Enrichr (https://maayanlab.cloud/Enrichr/). This involved performing functional enrichment analysis, specifically GO and KEGG pathway analysis, on all genes in the key modules. The analysis produced a list of the top 10 biological processes, cellular components, molecular functions and KEGG pathways, which were ranked based on their P-value. The results were then plotted to facilitate interpretation.

### 2.6. Using Cox model for identification of SRGs

The Cox model is a statistical method used in survival analysis to investigate the relationship between survival time and predictor variables. It allows for the estimation of hazard ratios to measure the relative risk of an event occurring in one group compared to another. In identifying hub genes, the Cox model assesses the association between gene expression and survival time of patients with a disease. By examining hazard ratios and p-values, it identifies genes strongly associated with survival, providing insights into disease processes. The Cox model has been applied to identify hub genes in diseases such as cancer and Alzheimer’s, aiding in understanding their underlying biology and developing new treatments. we applied Cox proportional hazards regression analysis to extract genes of 6 key modules that are strongly associated with survival outcomes and other clinical variables.

### 2.7. Using defective 3-parameter Gompertz model for identification of CSRGs

we employed the defective 3-parameter Gompertz model to analyze the impact of variables leading to risk-free subjects in 770 selected genes modules related to survival in RCC. The defective 3-parameter Gompertz model is a method for analyzing kidney cancer data and understanding the impact of various risk factors on the disease. By focusing on the identification of CSRGs, this model offers a unique perspective that has been largely overlooked in previous studies.

### 2.7. Hub genes identification

The genes selected modules, as well as the results of the Cox and defective Gompertz 3-parameter models, were uploaded separately into the STRING database. After filtering for a confidence value greater than 0.4, a protein-protein interaction (PPI) network was constructed. Three PPI networks were visualized using Cytoscape (v3.9.1). MCODE and the MCC and Degree algorithm of the CytoHubba app were used to identify the hub genes common to the three networks.

### 2.8. Estimation of cure rate of hub genes

The defective 3-parameter Gompertz distribution estimates the cure rate, which represents the proportion of patients who will not experience recurrence or death due to cancer. The minimum and maximum cure expression can help identify the optimal range of gene expression associated with better treatment outcomes, providing valuable insights for personalized treatment options. So, we calculated the minimum and maximum cure expression for identified hub genes.

The survival function of Gompertz 3-parameter distribution is as follows (Hajizadeh et al., 2023):

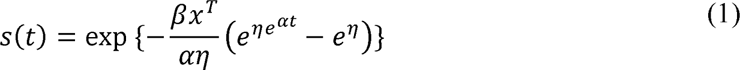

Here, α and η are shape parameters and belong to real values, *T* is survival time. x^T^f] = b_O_ + b_l_x+ b_2_x+ .. + b_k_x in which β = (b_0_, b_1, …,_ b_k_) indicates the coefficients vector to any gene and x^T^ = (1, x_O_, x_l_,…, x_k_) indicates the covariates vector which we enter any time one gene as covariate and achieved model as univariate. when α<0, we have the defective 3-parameter Gompertz (DG) model.

The cure fraction or cure rate of the DGD model is calculated based on the following formula(Hajizadeh et al., 2023):

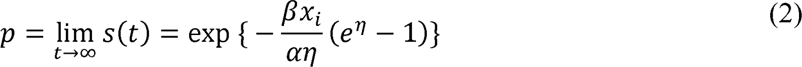

where p is the cure rate parameter.

### 2.9. AUC analysis calculation on hub genes

We used AUC analysis to evaluate the performance of hub genes in predicting kidney cancer outcomes using the SurvivalROC package and KM method of R software. AUC, or area under the curve, is a statistical measure that reflects the accuracy of a diagnostic test or predictive model. The higher AUC value, the more accurate the model is at distinguishing between positive and negative outcomes. In our study, we identified the top hub genes using a comprehensive network analysis approach and then performed AUC analysis on these genes to evaluate their prognostic power for kidney cancer prognosis.

### 2.10. Survival analysis of hub genes

The statistical analysis of overall survival (OS) performed using the “survival” package and event-free survival (EFS) performed using the programing in RStudio software (http://www.r-project.org/). To estimate the OS/EFS, Kaplan-Meier survival curves are utilized with the survft function, and the differences in OS/EFS between the high and low-risk groups are computed using a log-rank test and risk score determined by median. The data subjected to univariate analysis using the Cox proportional hazards regression model through the coxph function in R. The significance of the survival association determined by comparing the p-values of the log-rank and Wald tests with a threshold of 0.05.

## 3. Results

### 3.1. Identification of DEGs

Following data pre-processing and quality assessment, expression matrices were generated from the 22420 mRNAs. A stringent threshold of adj-P value < 0.05 and |log2FC| > 1 was applied to identify the DEGs. This resulted in a final set of 7940 DEGs, including 5182 up-regulated and 2758 down-regulated genes, which will be further analyzed in subsequent stages of the study (Figure. 3).

**Fig.3.**
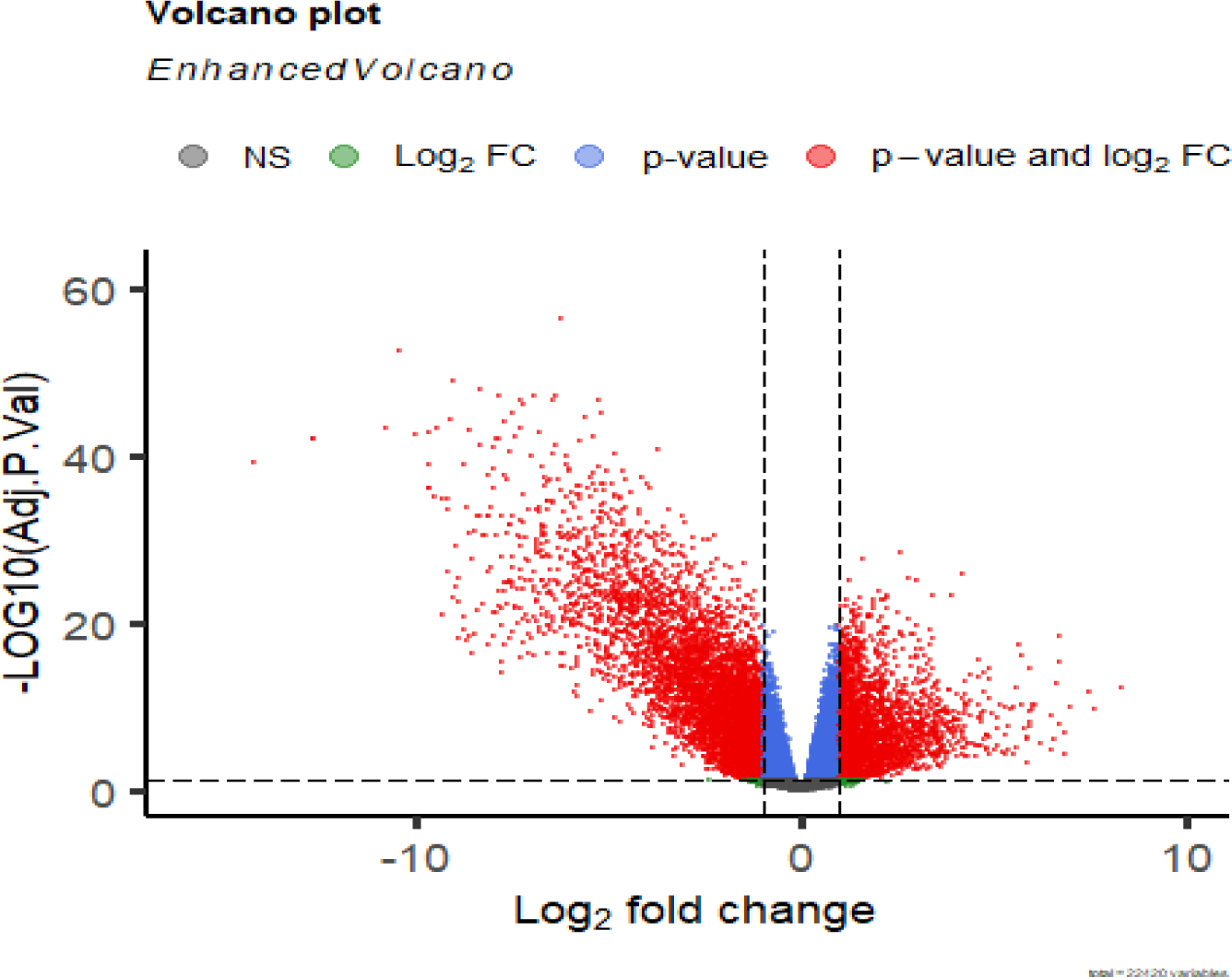
Visualizing Significant Differential Expression of Genes by Volcano Plot of DEGs.

### 3.2. Co-Expression Network construction

To find the correlation coefficients of the expression level in each of the 90 given samples, we clustered similar expression patterns of DEGs using the ’WGCNA’ package in R and removed one outlier sample (figure 4). We ensured a scale-free network by selecting a soft threshold of β=4 (with a scale-free R2 = 0.90) (figure 5). The gene dendrograms and the corresponding module colors of the identified modules are shown in Figure 6. Eighteen modules were identified, and we used two methods to test the association of each module with the KICH progression.

**Fig.4.**
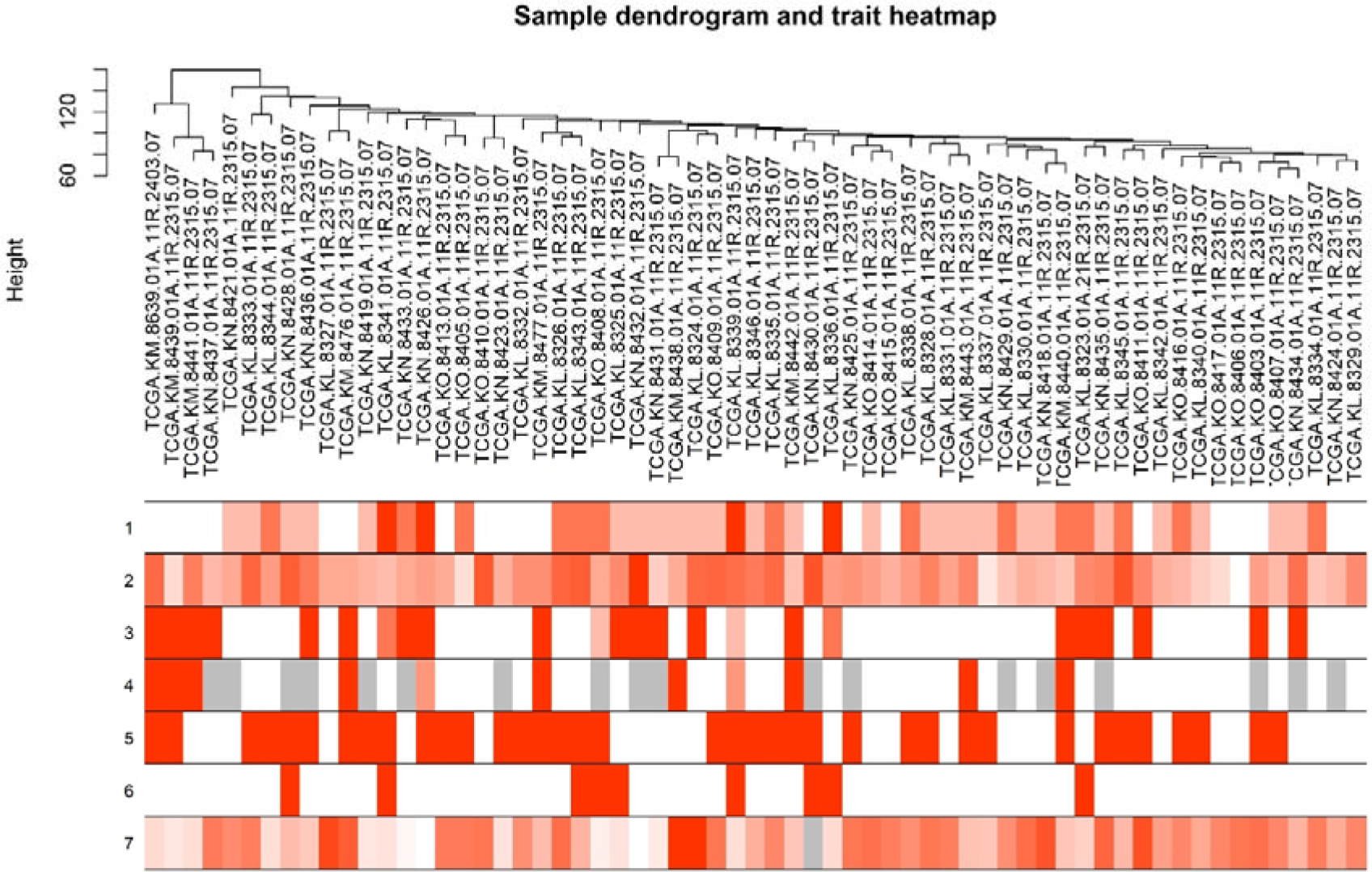
Dendrogram clustering of 90 KICH samples along with their clinical traits. Clustering based on expression data of genes differentially expressed between tumour and non-tumour KICH samples. Clinical traits (age, gender, cancer stage, pathology, metastasis, status, or overall survival (OS)) were proportional to color intensity.

**Fig.5.**
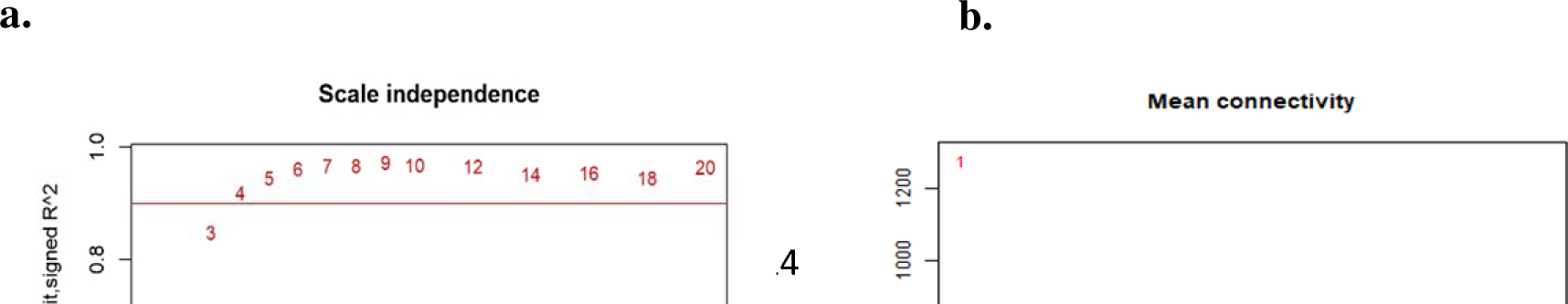
Highlights the process of determining the parameter β in the WGCNA algorithm for the adjacency function. a) analyzing the scale-free fit index for different soft thresholding powers, b) evaluating the mean connectivity for each thresholding power, c) checking the scale-free topology, and d) examining the connectivity distribution when β=4.

**Fig.6.**
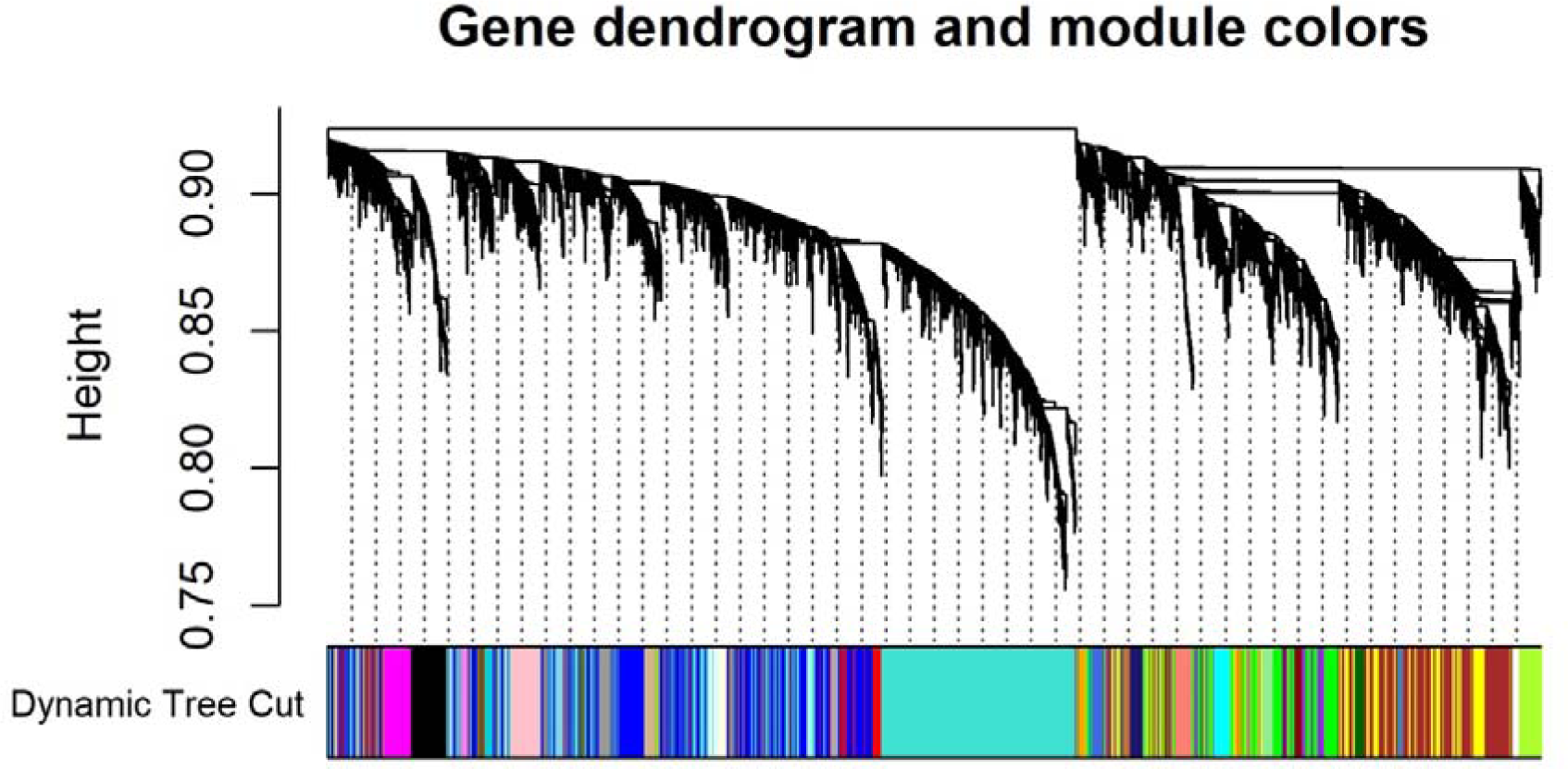
Dendrogram clustering of genes by hierarchical clustering adjacency-based dissimilarity.

### 3.3. Identifying the Clinically Significant Modules

Although most of the correlations were low to moderate (R^2^ < 0.90), we found that the 6 modules had a higher MS. Furthermore, the ME in the 6 modules (salmon, red, Eskyblue, darkgrey, black, and brown modules) showed a stronger relationship with cancer stage, pathology, metastasis, status and overall survival (OS). Therefore, we identified the 6 modules as the clinically significant module and extracted them for further analysis (figure 7). 770 genes with high connectivity in 6 key modules were identified as hub genes (figure 8).

**Fig.7.**
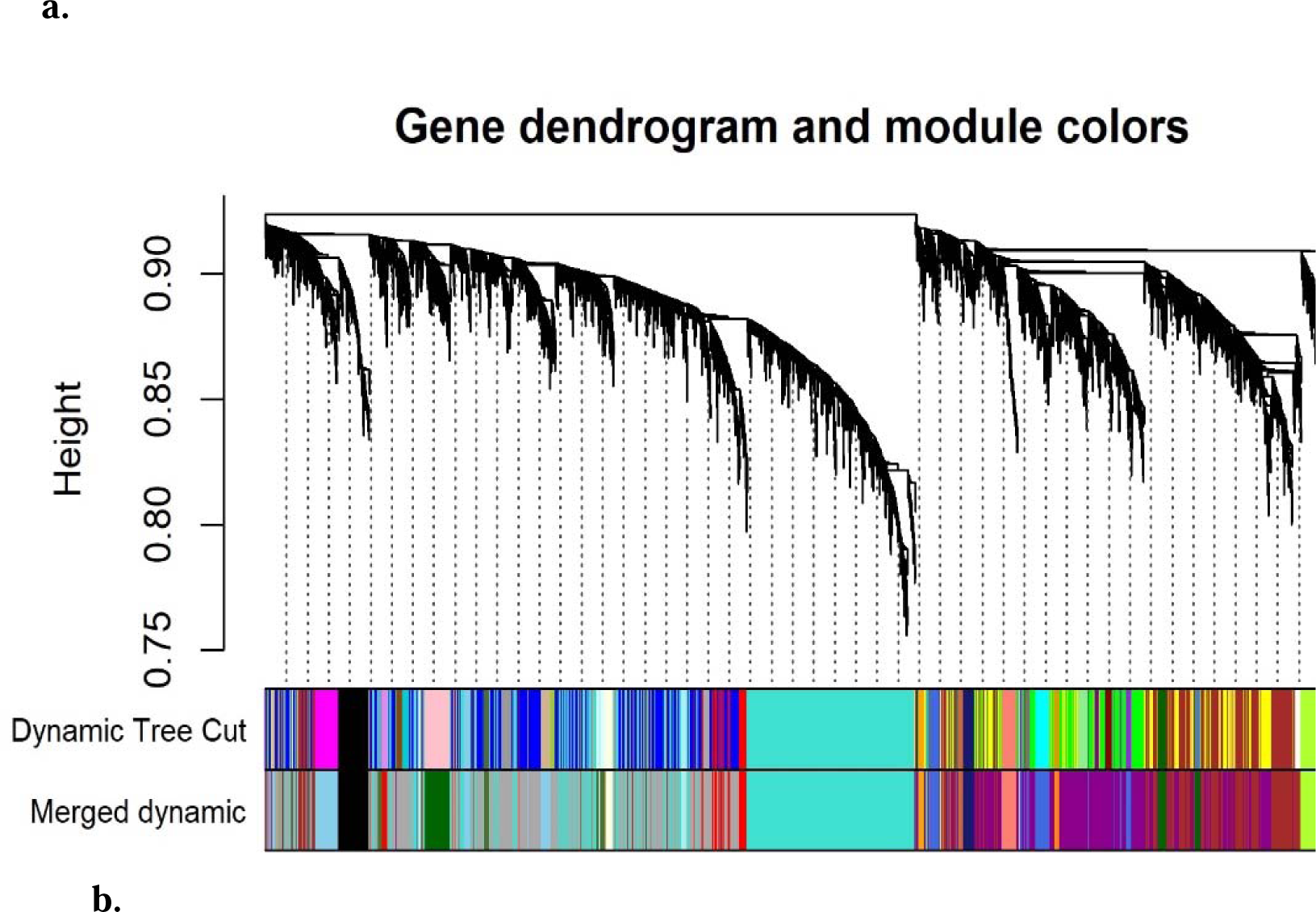
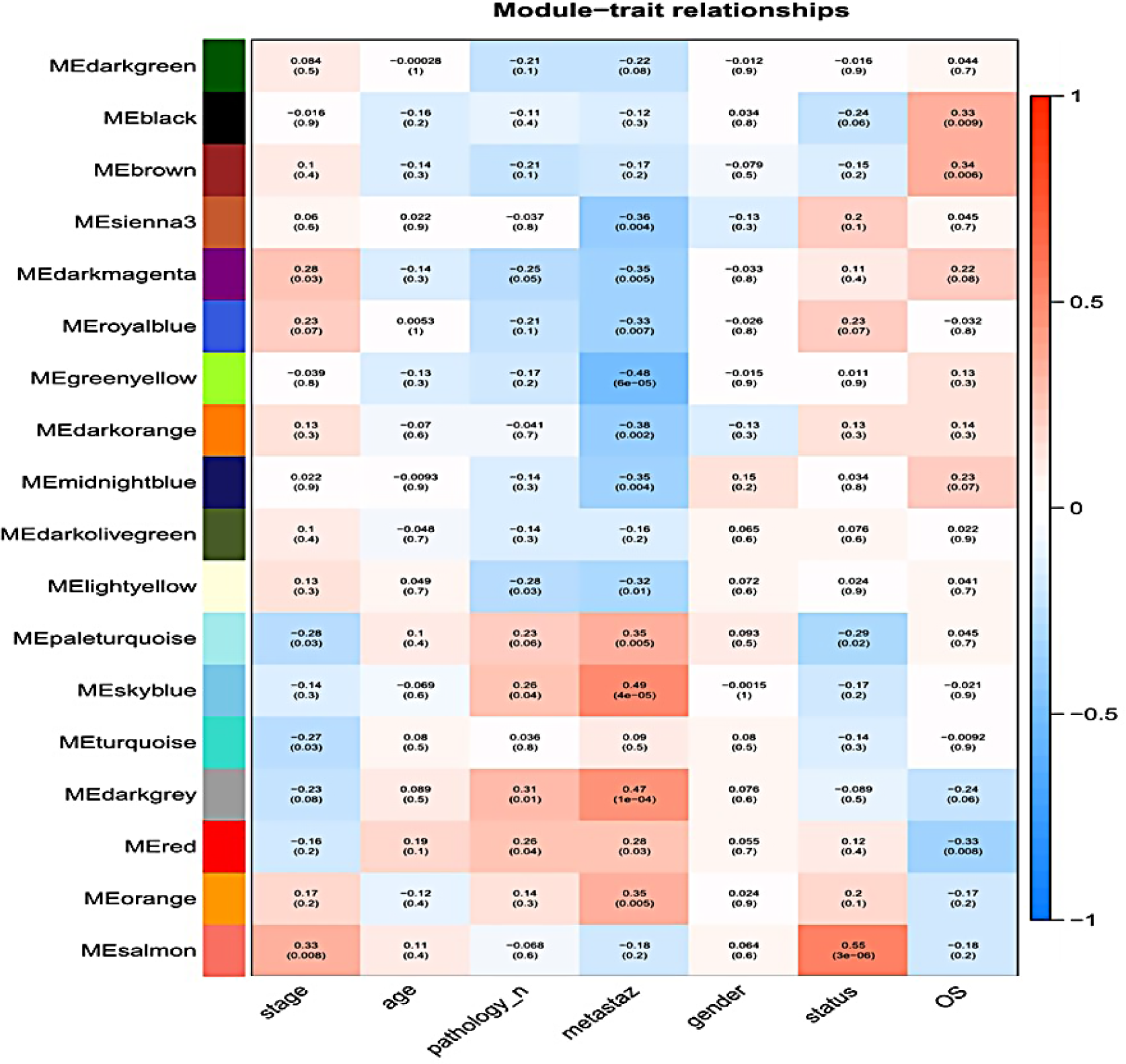
Identifying modules related to clinical traits. a) Dendrogram showing the clustering of all expressed genes that are different, using a measure of dissimilarity called “1-TOM”. b) Heatmap indicating the correlation between module eigengenes and various clinical features of kidney cancer, such as age, gender, cancer stage, pathology, metastasis, status or overall survival (OS) (Significant p-value≤0.05). In the heatmap, the color salmon represents the association between eigengenes and cancer stage and status, while red, Eskyblue, and darkgrey indicate the correlation with metastasis, and black and brown signify the relationship with overall survival.

**Fig.8.**
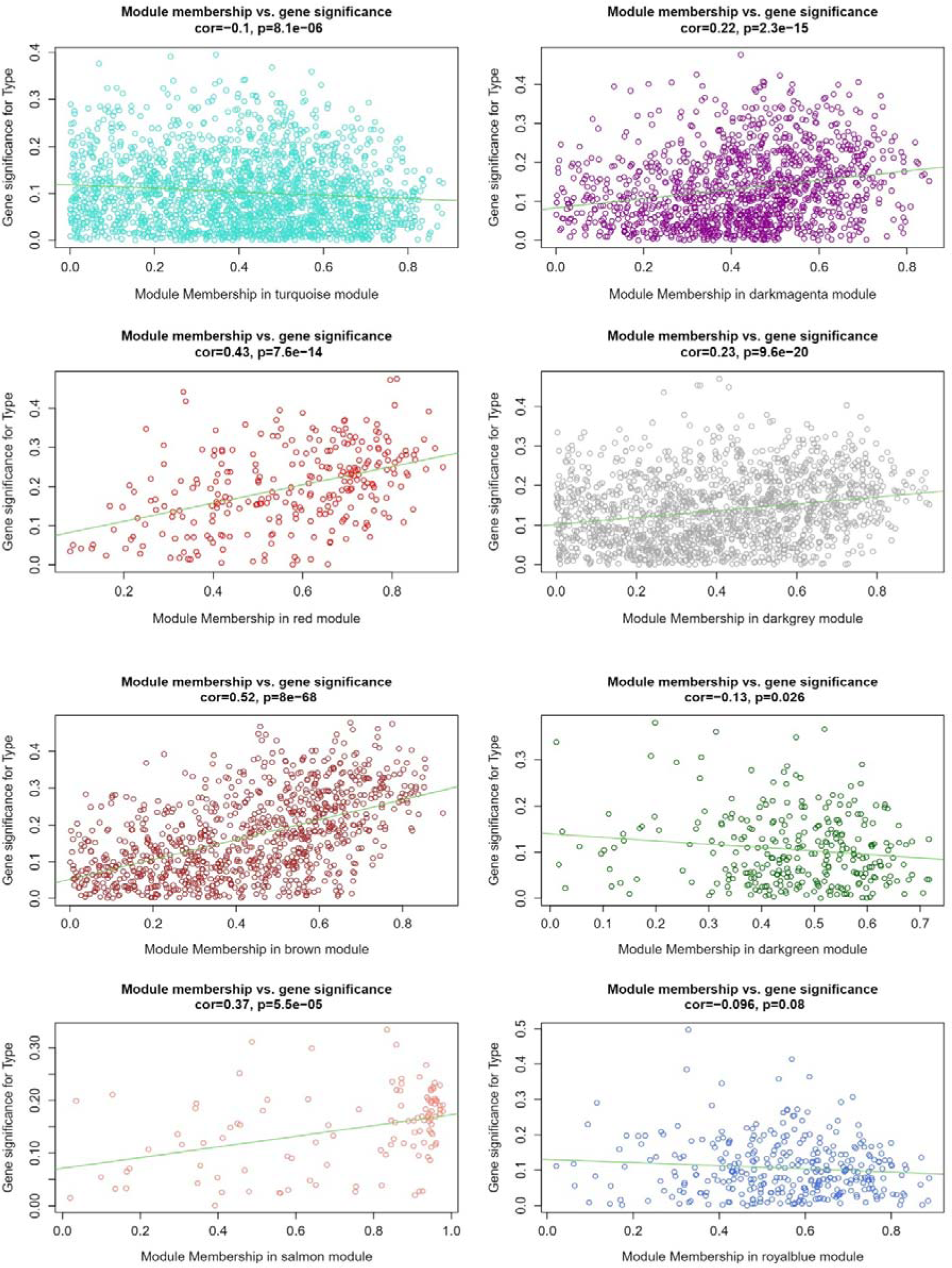
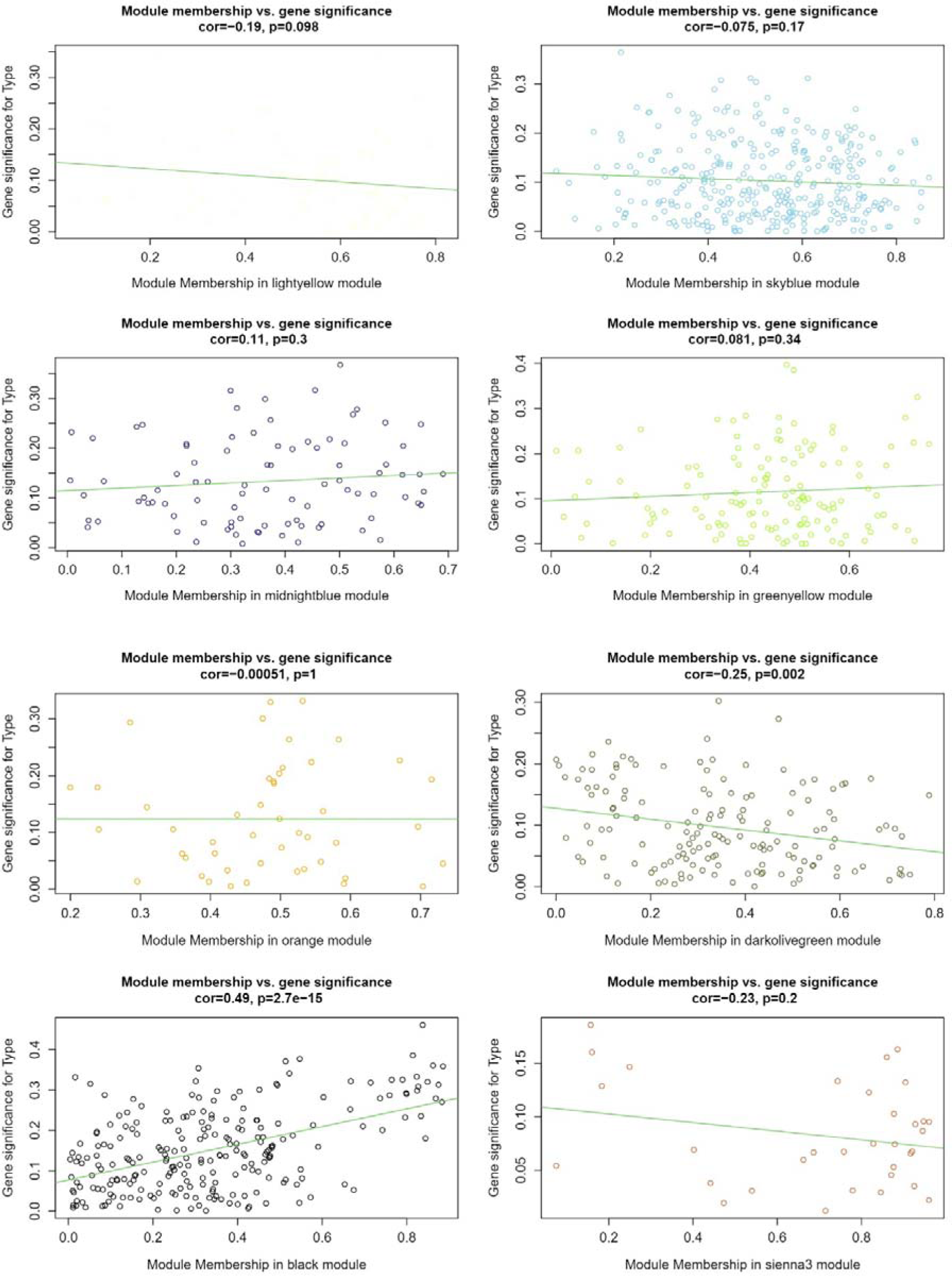
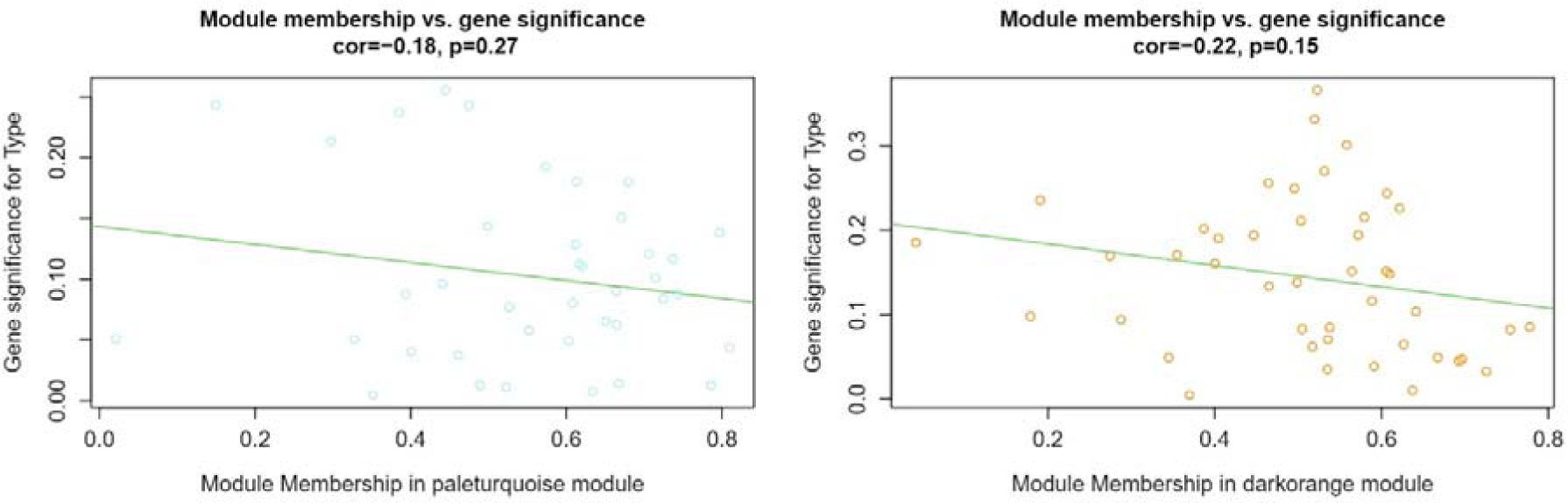
Module features of GS and MM. The relationship between the module’s membership of the significance of genes with clinical traits in all modules.

To identify the gene constitution of specific modules closely associated with overall survival, two unique features of the network - GS and MM - were utilized. Essentially, if the genes had highest absolute for both MM (0.7) and GS (0.75), they were considered particularly relevant to overall survival. Ultimately, 770 genes with high connectivity in 6 key modules were identified as hub genes. The red, brown, salmon, and black modules were found to have significant correlations between MM and GS, as shown in Figure 7. This suggests that these modules may play a particularly important role in the progression of kidney cancer.

### 3.4. Gene Pathway Analysis

As mentioned above, six modules were identified as significant and selected for further study. A total of 770 genes were extracted from these modules and analyzed using GO and KEGG pathway analysis via the Enrichr database to explore potential mechanisms. The results are shown in Figure 9 and indicate that the genes within the six modules are involved in a range of physiological processes and diseases, including Parkinson’s and Huntington’s diseases, as well as gastric acid secretion. The KEGG pathway analysis also indicated that these genes are involved in metabolic processes such as ascorbate and adorate metabolism and glyoxylate and dicarboxylate metabolism. Moreover, pathways related to bacterial infections such as Vibrio cholerae infection and Helicobacter pylori infection were also noted. These pathways suggest that bacterial infections may have a role in the progression or development of kidney cancer.

**Fig.9.**
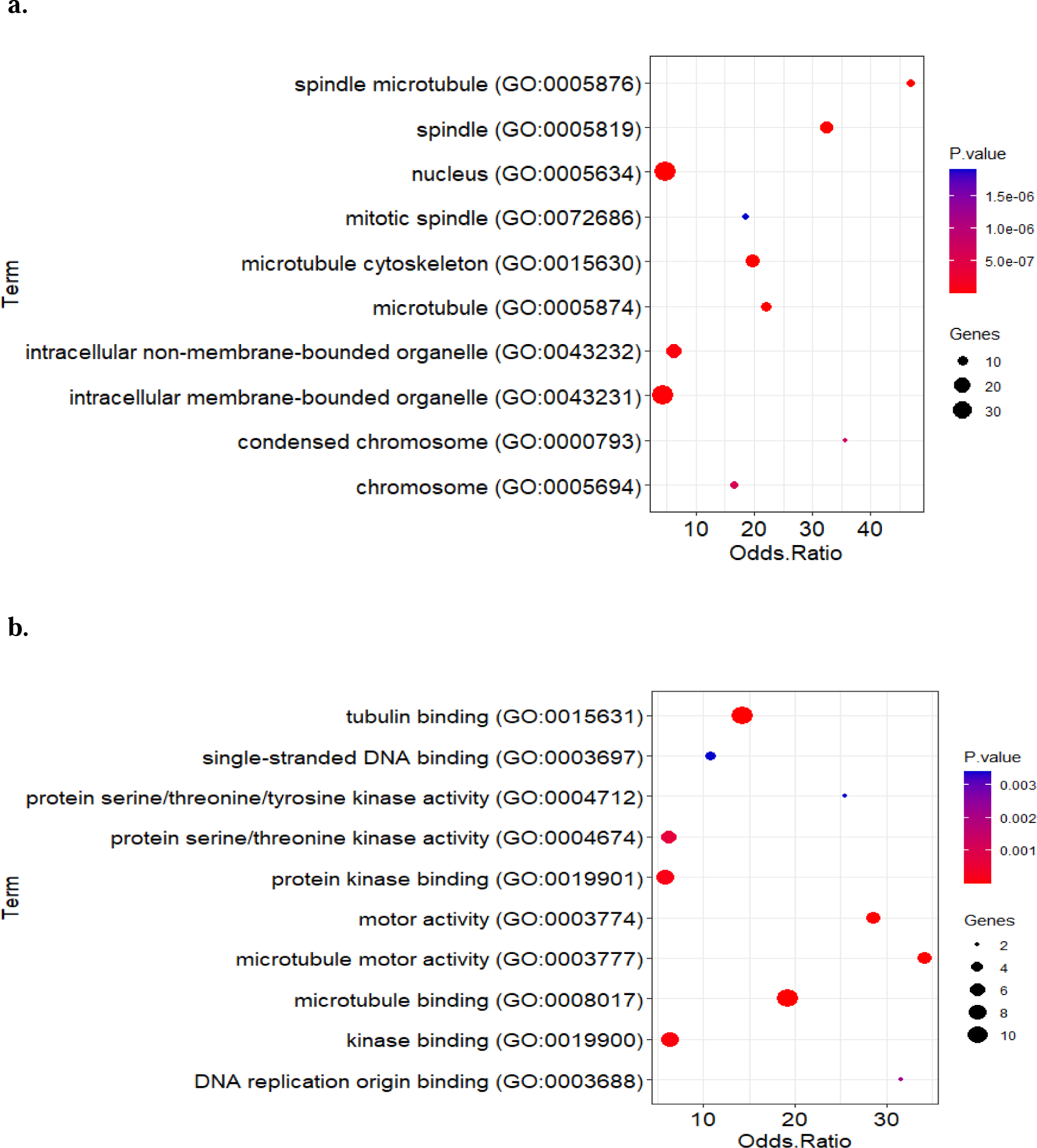
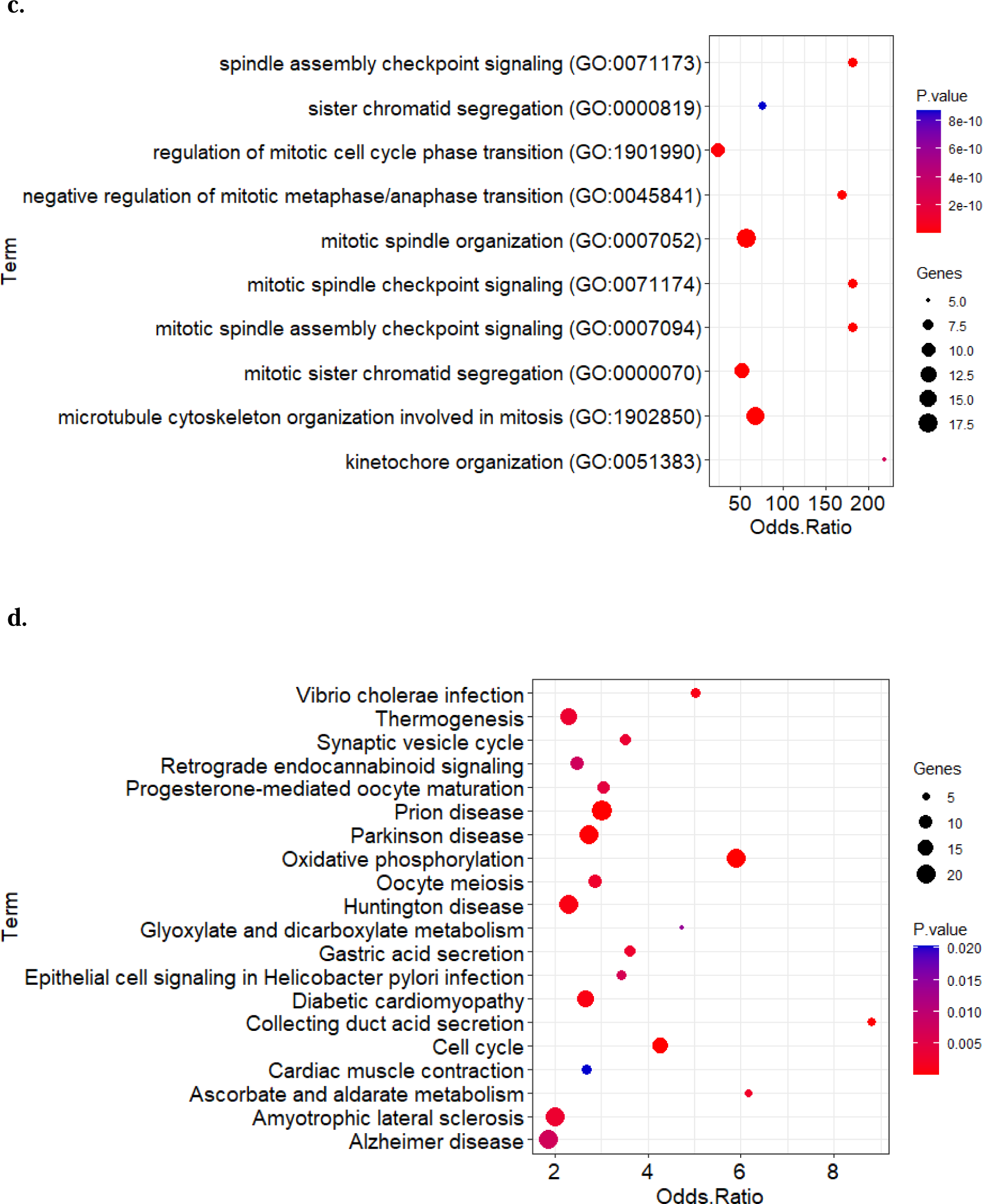
GO term and KEGG pathway analysis for genes of key modules. a) Cellular Component, b) Molecular Function, c) Biological Process and d) KEGG pathway enrichment analysis of key modules.

### 3.5. Outputs of Cox and 3-parameter defective Gompertz models

After analyzing 770 genes belonging to selected modules with both the Cox and 3-parameter defective Gompertz models separately, it was found that the output of both models was almost same, with only 6 genes showing variation between the two. Specifically, the Cox model identified 173 genes associated with kidney cancer, while the 3-parameter defective Gompertz model identified 175 genes. The Venn diagram analysis revealed that two genes, GFER and AL590096.1, were uniquely identified by the Cox model, while four genes, IFT81, RAI14, CSNK1E, and CLIC4, were unique to the 3-parameter defective Gompertz model (Figure 10) (Supplementary fig.1). However, the majority of the genes were common to both models.

**Fig.10.**
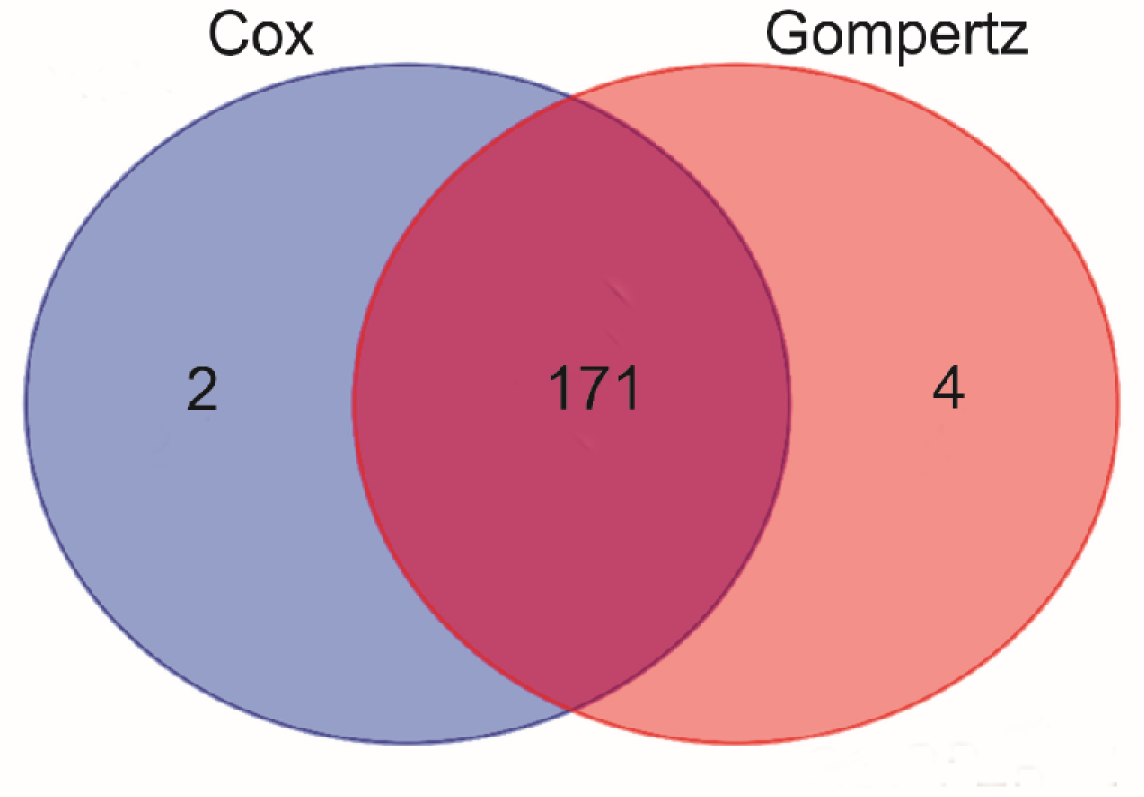
Venn diagram of the Cox/3-parameter defective Gompertz model output.

### 3.6. Identification of Hub genes in two PPI networks

In first, we examined the output of the Cox and 3-parameter defective Gompertz models (figure 11a, b) To construct the PPI network then by MCODE plugin that the best network was identified with 62 genes and a score of 57.9 (figure 11c). Also, we used network MCC and degree of CytoHubba app for the top 10 genes. To finding the hub genes present in two networks (figure 11d, e). Ultimately, we identified 10 hub genes (KIF20A, BUB1, AURKB, NCAPG, TOP2A, BUB1B, DLGAP5, TTK, TPX2, CDCA8) that were common in two networks. Meanwhile, 3 different hub genes (PLK1, CCNA2, CDC45) were observed in defective 3-parameter defective Gompertz and 4 different hub genes (PBK, CENPF, ASPM, CCNB2) in Cox PPI network (figure 12).

**Fig.11.**
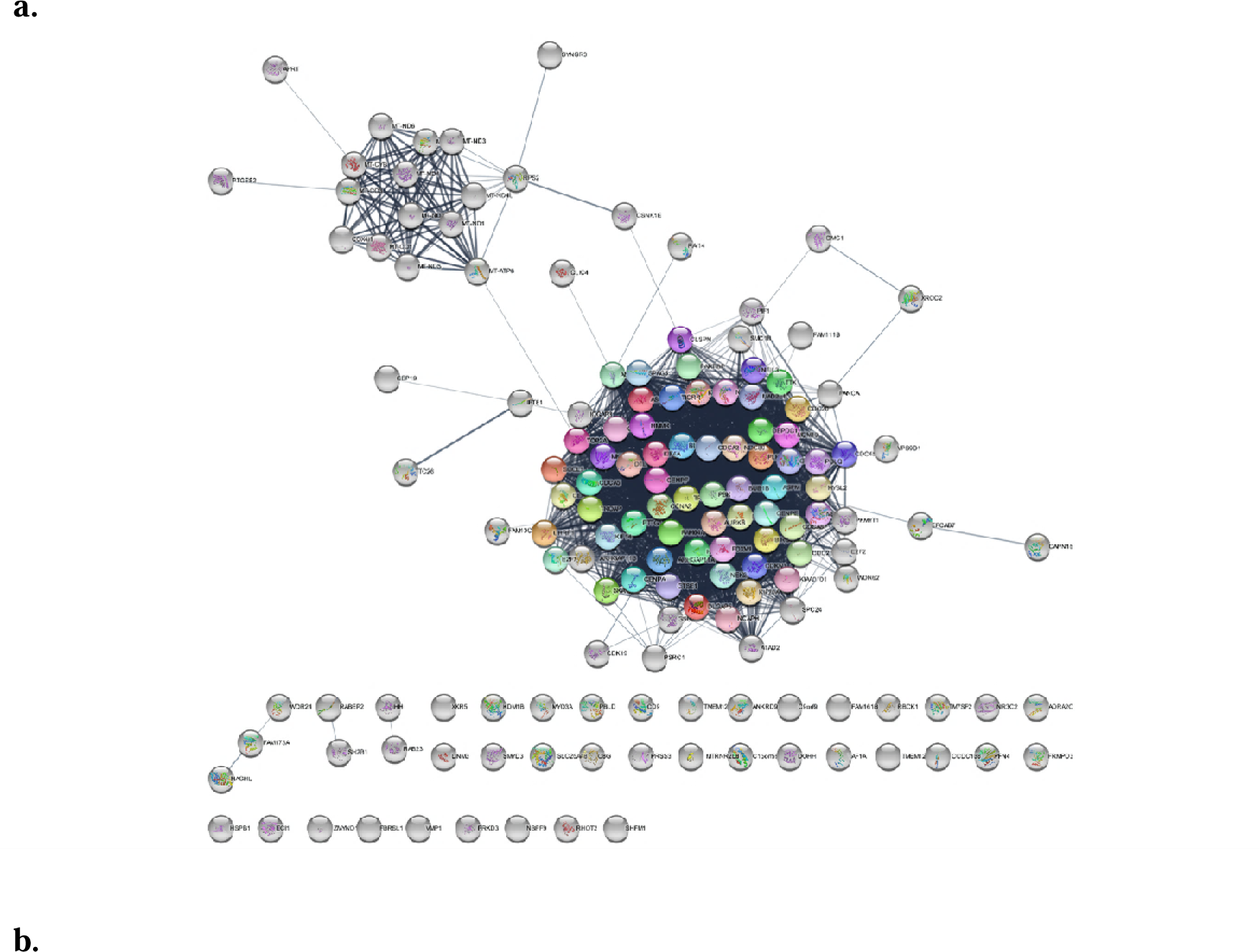
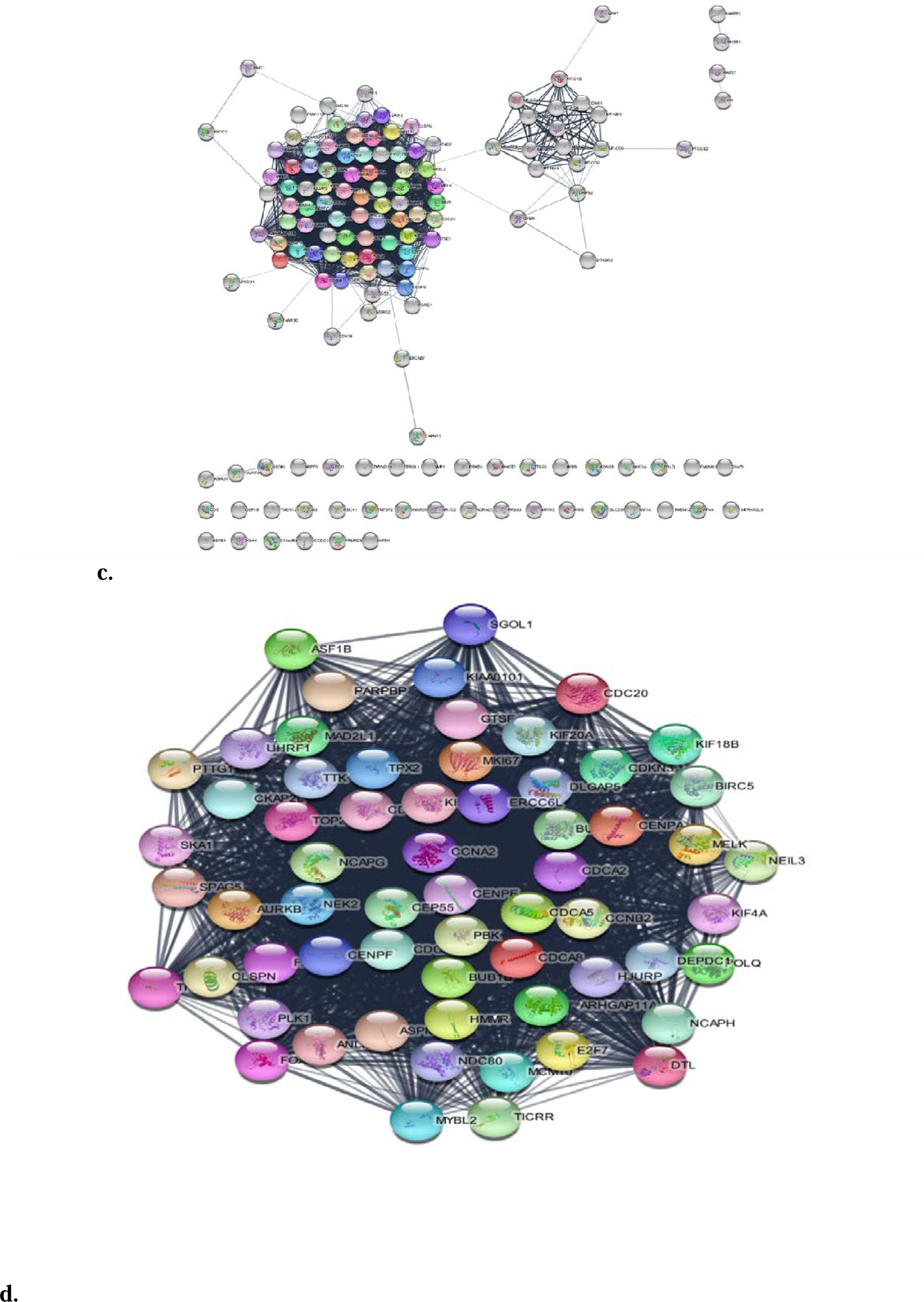
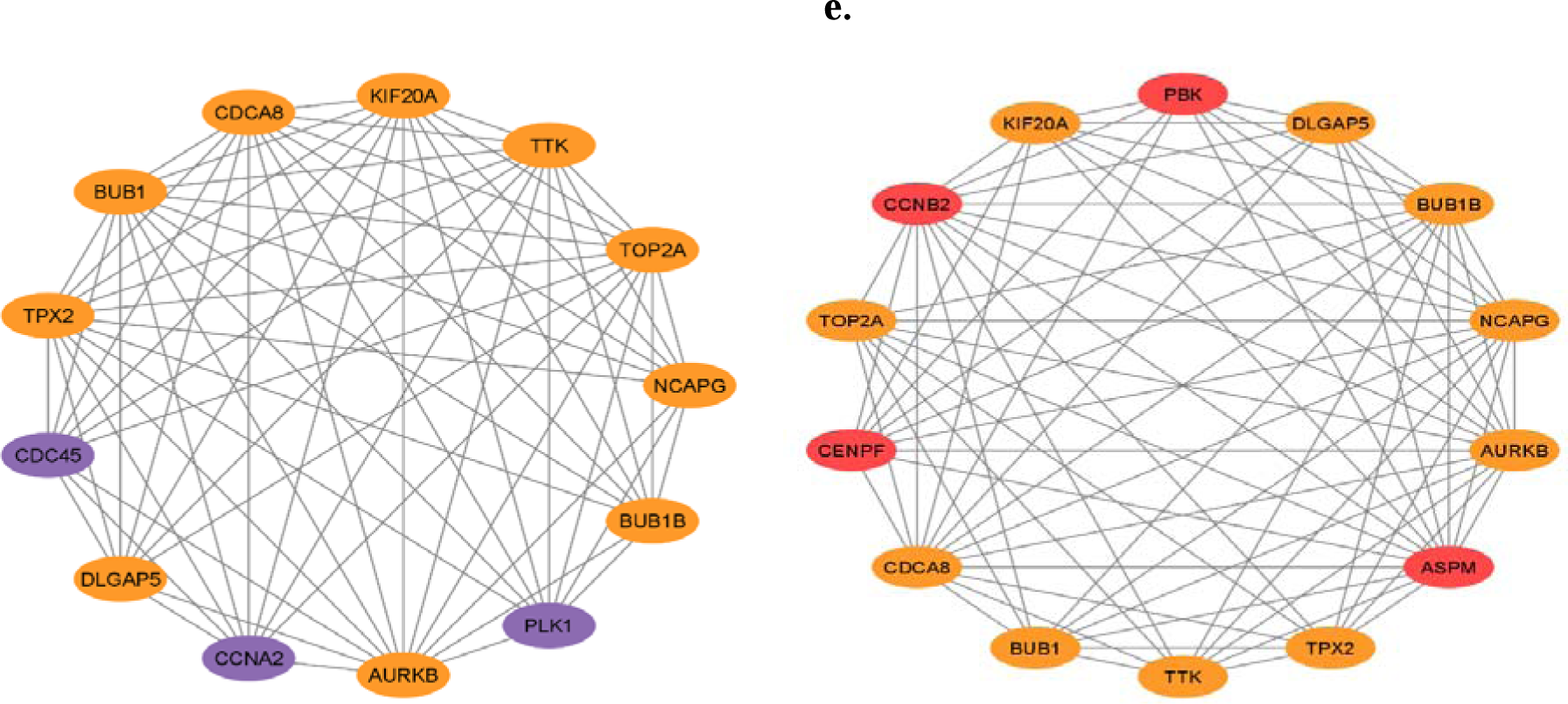
PPI network construction and screening for key genes. a) PPI network of defective Gompertz model output, b) PPI network of Cox model output c) MCODE cluster network of differentially expressed genes in cox and defective Gompertz output, d) merged networks of the top 10 MCC and the degree of the defective Gompertz model, e) merged networks of the top 10 MCC and the degree of the Cox model output. Note: Orange color = common hub genes, purple color = different hub genes with Cox model, red color = different hub genes with defective Gompertz model.

**Fig.12.**
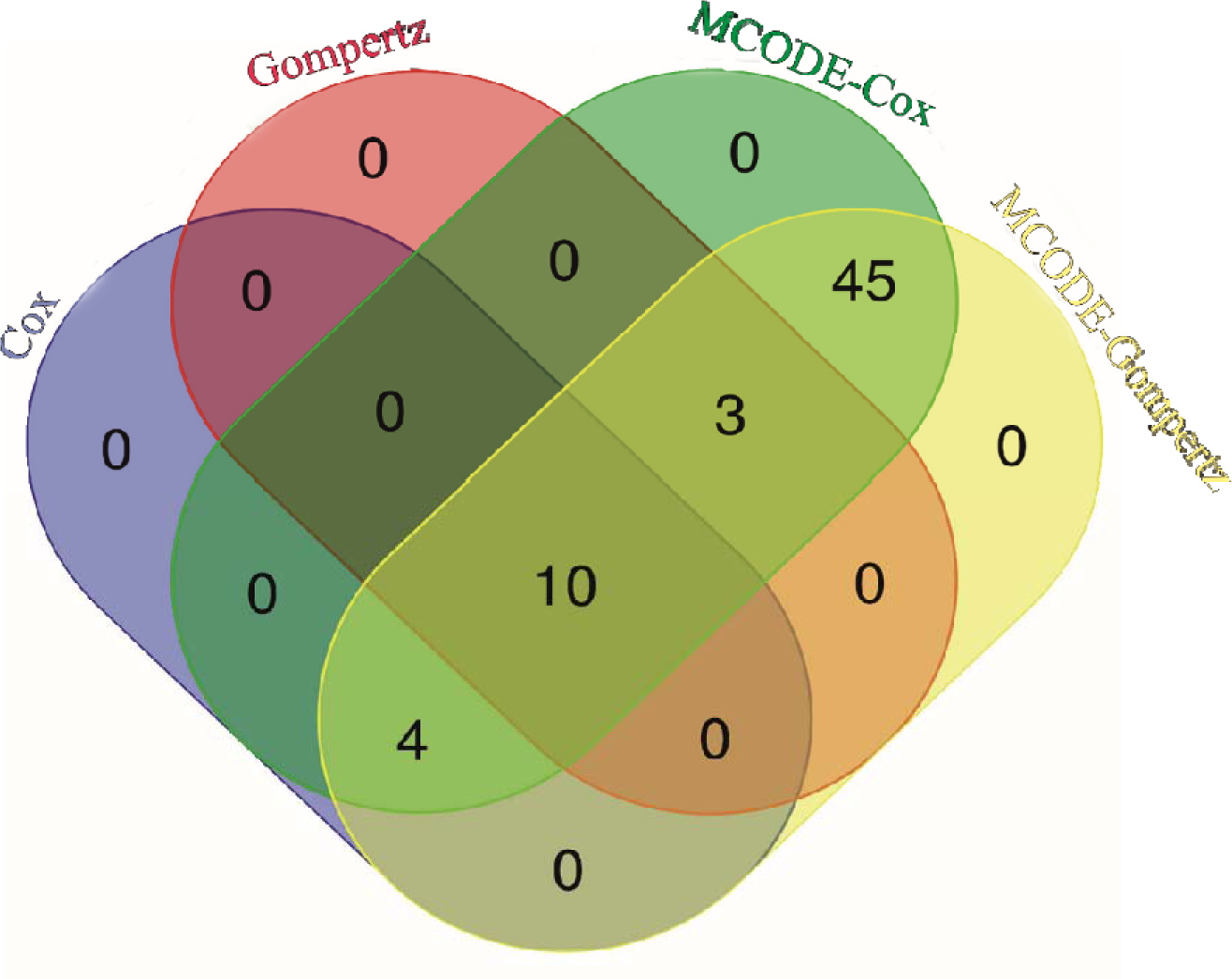
Venn diagram of two networks’ hub genes and MCODE cluster.

### 3.7. Cure rate estimation of hub genes

The findings of this analysis indicate a significant association between the expression levels of the identified hub genes and the cure rate in renal cell carcinoma. Specifically, the expression levels of six genes, namely TTK, KIF20A, DLGAP5, BUB1, AURKB, and CDC45, exhibited the strongest correlation with a reduction in the cure rate (table 1 and supplementary fig.1). These results suggest that targeting these genes may hold promise as a potential therapeutic strategy for improving treatment outcomes in RCC patients.

**Table 1.** AUC and min and max cure expression of common hub genes in Cox and Gompertz model.

| Gene name | Beta in cox | p-value | HR | Beta in Gompertz | p-value | Cure in min exp | Cure in max exp | HR | AUC |
| --- | --- | --- | --- | --- | --- | --- | --- | --- | --- |
| TPX2 | 0.792 | 0.000027 | 2.2086 | 0.784 | 0.000016 | 0.96 | 0.039 | 0.456462 | 0.781 |
| NCAPG | 1.083 | 0.000007 | 2.9537 | 1.077 | 0.0000028 | 0.955 | 0.034 | 0.340747 | 0.809 |
| TTK | 1.235 | 0.000002 | 3.4371 | 1.21 | 0.0000009 | 0.94 | 0.0005 | 0.447603 | 0.889 |
| KIF20A | 0.945 | 0.000009 | 2.5734 | 0.994 | 0.00000065 | 0.91 | 0.00006 | 0.370214 | 0.786 |
| DLGAP5 | 0.745 | 0.000023 | 2.1066 | 0.784 | 0.0000077 | 0.81 | 0.0000003 | 0.456549 | 0.806 |
| TOP2A | 0.711 | 0.000019 | 2.0366 | 0.704 | 0.000011 | 0.96 | 0.034 | 0.494858 | 0.808 |
| CDCA8 | 0.805 | 0.000044 | 2.2359 | 0.795 | 0.0000303 | 0.93 | 0.045 | 0.451452 | 0.762 |
| BUB1B | 0.91 | 0.000033 | 2.4793 | 0.961 | 0.0000131 | 0.95 | 0.05 | 0.38265 | 0.830 |
| BUB1 | 0.834 | 0.0000448 | 2.3030 | 0.82 | 0.0000327 | 0.92 | 0.003 | 0.440453 | 0.857 |
| AURKB | 1.081 | 0.00000516 | 2.9475 | 1.013 | 0.000000156 | 0.93 | 0.0000001 | 0.362972 | 0.77 |

### 3.8. Prognostic value of hub genes

In this study, we identified a total of 14 hub genes in the Cox model and 13 hub genes in the Gompertz model, of which 10 hub genes were common between the two models. We assessed the prognostic value of these hub genes using the survivalROC package and KM method, with a cutoff point of 4676 days (12 years) for calculating gene expression in individuals’ follow-up time. The results showed that almost all hub genes had an AUC value higher than 0.93 for one year, indicating excellent prognostic value. Additionally, hub genes with AUC values between 0.7 and 0.89 exhibited good diagnostic accuracy, indicating their potential use as diagnostic biomarkers (Table 1-3). According to our analysis, among the hub genes identified, six genes (NCAPG, TTK, DLGAP5, TOP2A, BUB1B, and BUB1) had the highest area under the curve (AUC) values.

### 3.9. km-plots of hub genes

The findings from the survival analysis of hub genes indicate a significant association between gene expression and overall survival (OS) in renal cell carcinoma. The use of Kaplan-Meier survival curves and log-rank tests enabled the computation of differences in OS between the high- and low-risk groups. Notably, in all survival curves, the graph appeared as a straight line (except TPX2), indicating cure (figure 13).

**Fig.13.**
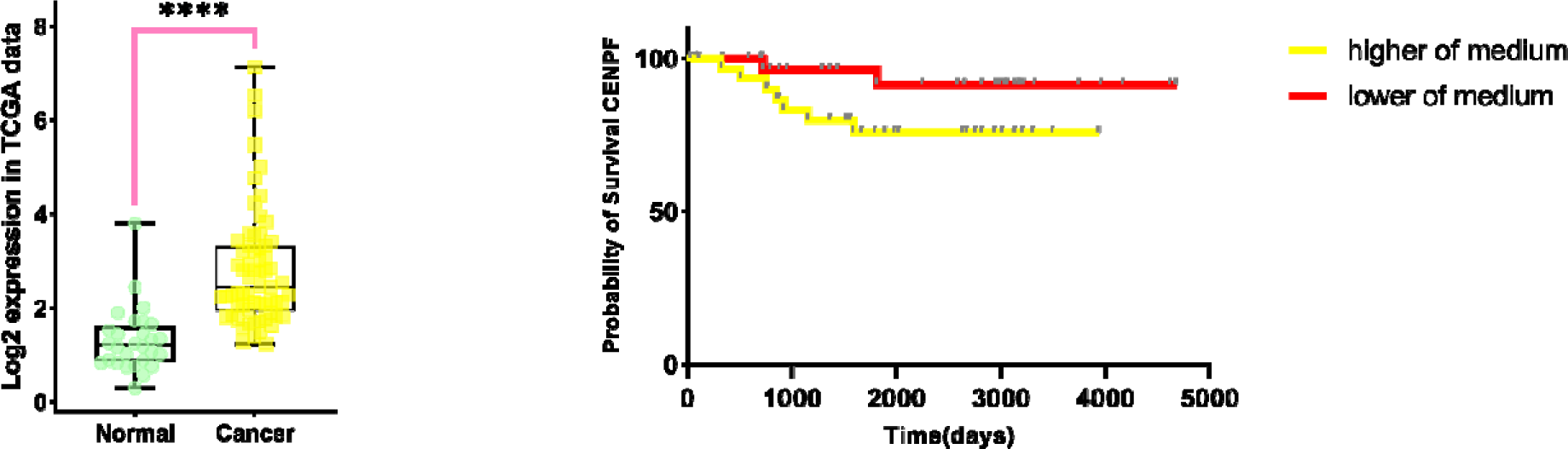
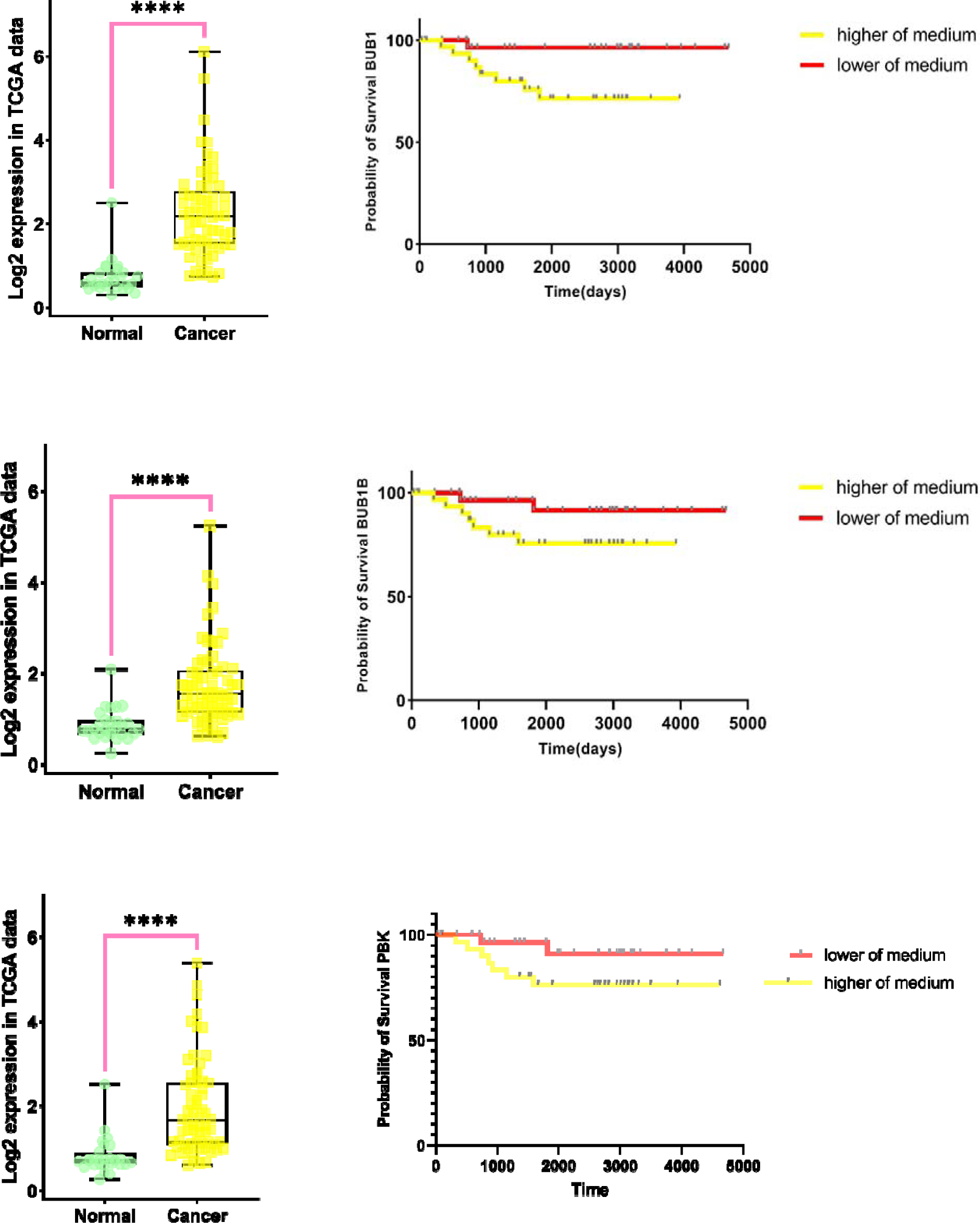
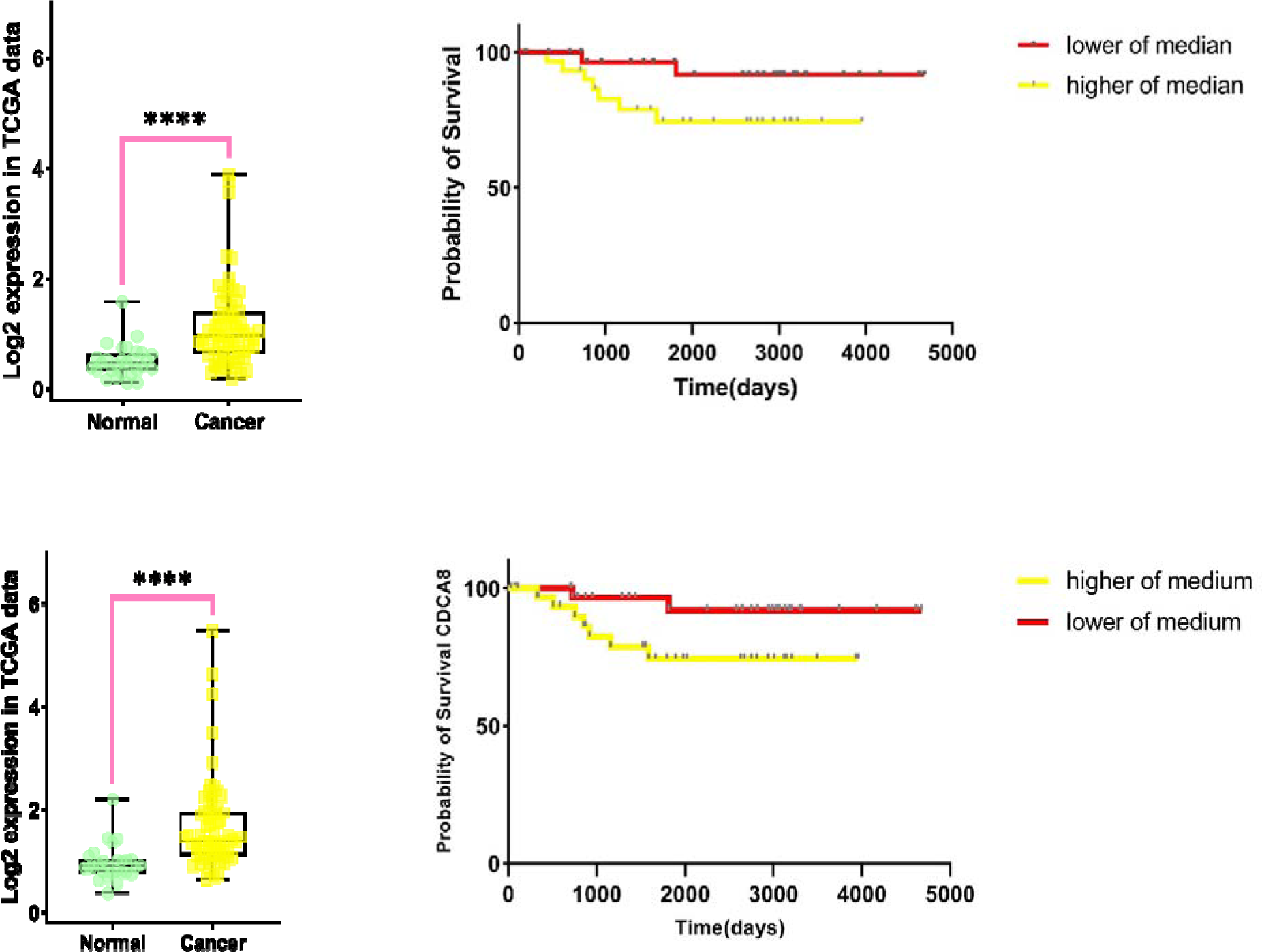
Box plots (left) and km-plots (right) of hub genes In the box plots, the significance levels were denoted as ****, indicating a p-value of ≤0·00001, and ** representing a p-value of ≤0·001. In the Kaplan-Meier (KM) plots, all lines are represented by a long, flat tail that is noticeably distant from zero. This pattern suggests a significant survival advantage or potential for cure in kidney cancer patients.

## 4. Discussion

After extraction mRNA expression profiles of RCC cancer and data normalization, six key modules related to cancer stage, pathology, metastasis, status and OS (fig.7 were identified by WGCNA technique and key genes were selected based on their Module Membership (MM) and Gene Significance values. Key pathways and key genes were identified via KEGG pathway enrichment analysis and PPI network method respectively. Also, SRGs and SCRGs were identified by Cox proportional hazards model and the 3-parameters defective Gompertz model and observed differences in the results obtained from both statistical methods. The findings revealed that the hub genes identified in these two models were not entirely identical. Specifically, the 3-parameters defective Gompertz model identified three different hub genes, namely PLK1, CCNA2, and CDC45, while the Cox PPI network identified four different hub genes, namely PBK, CENPF, ASPM, and CCNB2.

PLK1, CCNA2, and CDC45 are all involved in cell cycle regulation and have been implicated in the development and progression of various cancers(Gheghiani et al., 2021, Iliaki et al., 2021, Matthews et al., 2022). PLK1 is a Ser/Thr kinase that is expressed predominantly during the G2/S and M phase of the cell cycle and is frequently overexpressed in various human cancers(Gheghiani et al., 2021, Iliaki et al., 2021). CCNA2 is a cyclin that regulates the G1/S transition of the cell cycle and is overexpressed in many types of cancer(Jiang et al., 2022). CDC45 is a protein that is involved in DNA replication and is overexpressed in several types of cancer(Moyer et al., 2006, Srinivasan et al., 2013). Additionally, these genes, as reported in Table 2, are highly relevant to the cure rate of RCC patients, as their maximum expression strongly reduces the cure rate.

**Table 2.** AUC and min and max cure expression of Gompertz hub genes.

| Gene name | Beta | p-value | Median in expression | Cure in min | Cure in max | HR | AUC |
| --- | --- | --- | --- | --- | --- | --- | --- |
| CDC45 | 1.235 | 0.000003 | 0.960447 | 0.84 | 0.0000034 | 0.29067 | 0.774 |
| CCNA2 | 0.847 | 0.000132 | 1.599271 | 0.95 | 0.10 | 0.428625 | 0.778 |
| PLK1 | 0.855 | 0.0000228 | 1.319569 | 0.94 | 0.057 | 0.425247 | 0.736 |

**Table 3.** AUC of Cox hub genes.

| Gene name | Beta | p-value | Median in expression | AUC | HR |
| --- | --- | --- | --- | --- | --- |
| PBK | 0.815676 | 0.000038 | 1.366824 | 0.797 | 2.26 |
| CENPF | 0.756063 | 0.0000177 | 2.380264 | 0.786 | 2.12 |
| ASPM | 0.765976 | 0.000023 | 1.168842 | 0.783 | 2.15 |
| CCNB2 | 0.81976 | 0.000066 | 1.813181 | 0.787 | 2.27 |

PBK, CENPF, ASPM, and CCNB2 have also been implicated in cancer development and progression. PBK is a serine/threonine kinase that is overexpressed in many types of cancer, including kidney cancer, and is associated with poor prognosis(Nagano-Matsuo et al., 2021, Wen et al., 2021). CENPF is involved in cell division(Varis et al., 2006). Also, CENPF is involved in the centromere-kinetochore complex and has been found to impact cell proliferation and metastasis in multiple types of cancer, including kidney cancer(He et al., 2020, Huang et al., 2021b, Li et al., 2021). ASPM is a microtubule-associated protein that is involved in mitotic spindle assembly, and its overexpression has been observed in many cancers, including kidney cancer. CCNB2 is a regulatory subunit of CDK1, which plays a critical role in mitosis, and has been shown to be overexpressed in various cancers.

As reported in the results section, the expression levels of TTK, KIF20A, DLGAP5, BUB1, AURKB, and CDC45 demonstrated a significant association with decreased cure rate in renal cell carcinoma(Wan et al., 2019). Notably, the highest expression levels of AURKB and DLGAP5 were associated with the lowest chance of cure, suggesting that the overexpression of these two genes may lead to a reduced likelihood of successful treatment. AURKB and DLGAP5 are involved in multiple pathways that have been shown to be critical in the development and progression of kidney cancer.

AURKB has been identified as a promising biomarker in clear cell renal carcinoma and has been found to play a key role in the tumorigenesis and progression of renal cell carcinoma (Fang et al.). The AURKB gene is involved in the regulation of the cell cycle and is essential for the accurate segregation of chromosomes during cell division. Dysregulation in expression of AURKB has been implicated in the development of various cancers, including lung cancer, colorectal cancer, prostate cancer, breast cancer, and liver cancer (Pohl et al., 2011, Tang et al., 2017, Fang et al., 2018, Marima et al., 2021). AURKB has emerged as an attractive drug target leading to the development of small molecule inhibitors. Inhibition of AURKB activity has been shown to disrupt development and maintain proliferation and differentiation at later stages of development(Shaalan et al., 2021). However, it should be noted that AURKB gene expression levels were significantly up-regulated in kidney cancer patients compared to normal tissues.

DLGAP5 is a protein that is phosphorylated by AURKA, and it is one of its direct targets. DLGAP5 is involved in the formation of tubulin polymers which lead to tubulin sheets surrounding the microtubules. This protein plays an essential role in mitotic spindle formation during cell division. The phosphorylation of DLGAP5 by AURKA helps in stabilizing its association with the mitotic spindle(Schneider et al., 2017). DLGAP5 has been found to be overexpressed in several cancers, including breast, ovarian, colorectal, and lung cancers, and is often associated with poor prognosis and disease progression. Also, its high area under the curve (AUC) value of 0.8 suggests its potential as a prognostic biomarker for kidney cancer.

Five genes (NCAPG, TTK, TOP2A, BUB1B, and BUB1) besides DLGAP5, have an area under the curve (AUC) above 0.8, indicating their potential as prognostic biomarkers for RCC. These genes are involved in critical cellular processes, such as cell division, DNA replication, and chromosomal segregation, making them potential targets for cancer therapy. For instance, TOP2A encodes a topoisomerase that plays a crucial role in DNA replication and chromosome segregation during mitosis, and its overexpression has been observed in various types of cancer (Uusküla-Reimand and Wilson, 2022). Similarly, BUB1 and BUB1B encode kinases that are involved in spindle checkpoint function, and their dysregulation has been implicated in the development and progression of cancer (Jiang et al., 2021, Sekino et al., 2021). NCAPG and TTK are both involved in cell cycle regulation and have been found to be overexpressed in various cancers, including renal cell carcinoma (Li et al., 2022, Liu et al., 2019, Zhang et al., 2018). The identification of these genes as potential biomarkers can aid in the development of personalized treatment strategies for cancer patients.

in the end, it can be said that cure models such as defective models have better diagnostic power in identifying the effective factors in survival time than common survival models such as Cox proportional hazard model due to the fact that cure fraction in KM curve and long time follow.

## 5. Conclusion

In conclusion, the defective 3-parameters Gompertz model offers several advantages over the Cox proportional hazards model when analyzing mRNA expression profiles in cancer types such as renal cell carcinoma. One of the main benefits of the Gompertz model is that it provides more comprehensive information about the expression of survival-related genes, including the calculation of the minimum and maximum expression of cure rate. In contrast, the Cox model only calculates hazard ratios, which may not fully capture the complexity of gene expression patterns in cancer. This additional information is especially valuable in identifying potential therapeutic targets that may not be evident using the Cox model alone. Additionally, the Gompertz model’s ability to identify survival-related genes (SRGs) that are not captured by the Cox model may lead to the discovery of new therapeutic targets for RCC. Overall, the Gompertz model’s comprehensive approach to analyzing mRNA expression profiles in RCC provides a more nuanced understanding of the disease’s molecular mechanisms and offers potential avenues for improving patient outcomes.

### CRediT authorship contribution statement

**Maryam Ahmadian**: Conceptualization, Methodology, Software, Knowledge-based design, Validation, Formal analysis, Resources, Writing – original draft, Writing – review & editing, Visualization, Project administration. **Zahra Molavi**: Conceptualization, Knowledge-based design, Validation, Resources, Writing – original draft, Writing – review & editing, Visualization. **Ahmad Reza:** Conceptualization, Methodology, Software, Knowledge-based design, Validation, Formal analysis, Supervision, Funding acquisition. **Ali Akbar Maboudi**: Methodology, Project administration.

## Data Availability

All data produced are available online at:
Cancer Genome Atlas (TCGA) database (https://portal.gdc.cancer.gov/)

## Acknowledgements

Not applicable.

## Ethical approval

Non-applicant.

