## supplemental for "Identification Cure Hub Genes of Chromophobe Cell Renal Carcinoma : A study based on Weighted Gene Co-expression Network Analysis (WGCNA) and the Cure Defective Models"

Supplementary fig.1. Analysis of gene clusters. The branches in the dendrogram represent groups of clustered samples.

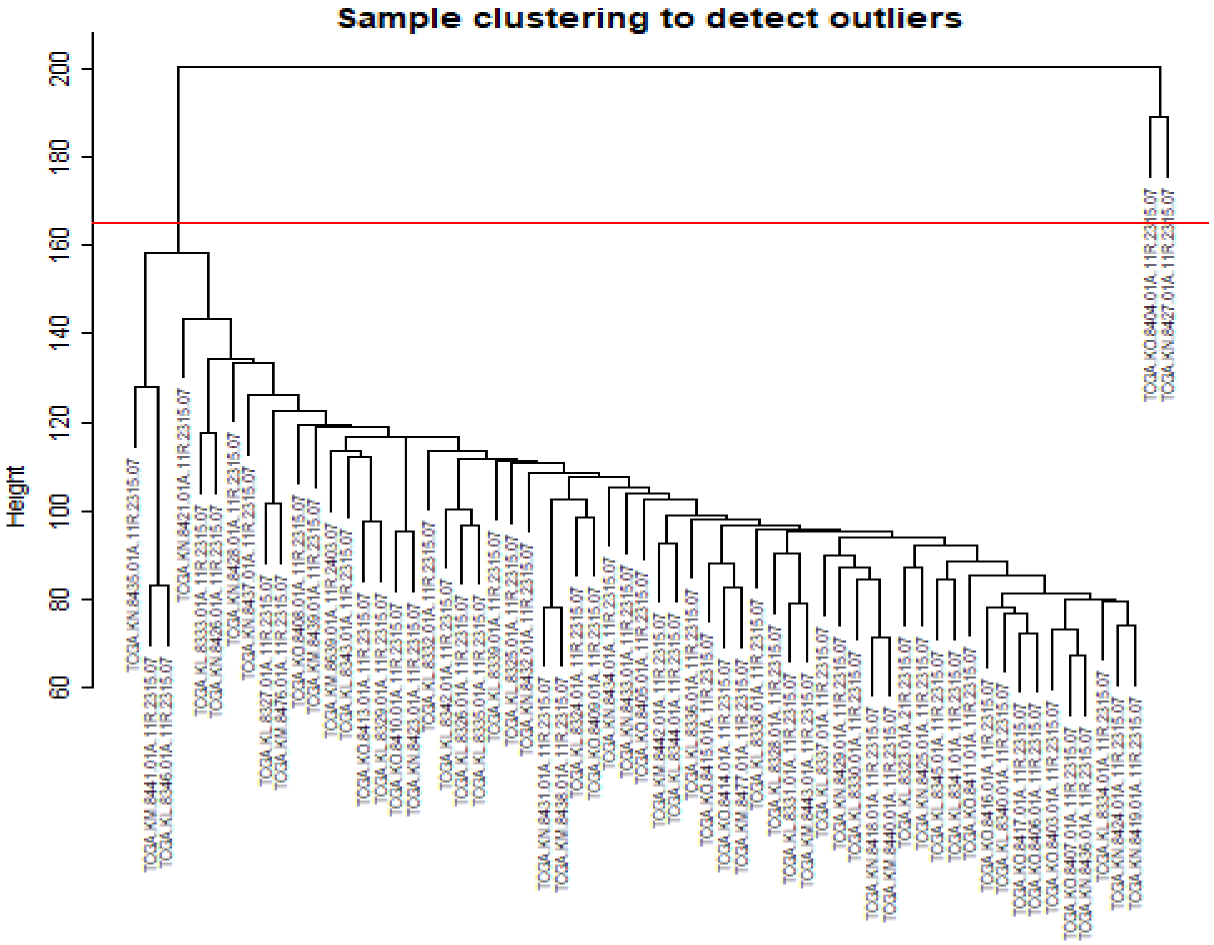


Supplementary table.1. Different output genes' cure rate expression in Compertz model with Cox model

| | Gene_name | Beta | P_Value | Cure Min exp | Cure Max exp | HR |
|---|---|---|---|---|---|---|
| ENSG00000213923.13 | CSNK1E | -1.386 | 0.0478 | 0.68 | 0.91 | 3.998689 |
| ENSG00000039560.14 | RAI14 | 0.933 | 0.0460 | 0.94 | 0.28 | 0.393403 |
| ENSG00000122970.16 | IFT81 | 0.855 | 0.000022 | 0.91 | 0.70 | 0.304178 |
| ENSG00000169504.15 | CLIC4 | 0.760 | 0.0486 | 0.90 | 0.67 | 0.467447 |
